# Efficacy of Rezūm® in reducing prostate volume. A retrospective study in patients with benign hyperplasia in Costa Rica

**DOI:** 10.1101/2024.12.22.24319507

**Authors:** Mario González, Dra. Milena Castro

## Abstract

**Aim:** To estimate the effect size of the prostate volume reduction after application of water vapor injections (Rezūm®) in 40 years or older patients, that have been diagnosed with Benign Prostate Hyperplasia (BPH) and are registered at UNIBE Hospital. This contrast provides a retrospective efficacy of the minimally invasive treatment valid for the local population.

**Methods:** A retrospective study was conducted to review patient records, attended at UNIBE Hospital, in Costa Rica. Prostate volume was established as the main variable to assess BPH reversion. Other variables like the International Prostate Symptome Score (IPSS) and urinary flow rate were also evaluated for each patient. Two sample mean comparisons of baseline and follow up measures for prostate volume and IPSS were calculated to estimate a mean difference. Effect size parameters like Cohen’s *d*, Hedges’s *g* and Glass’s *δ* were also estimated and compared based on the standard deviation assessment. A linear regression model was also adjusted to obtain values of the statistical contribution from each of the variables included to evaluate the clinical reversal of a benign prostate hyperplasia diagnosis.

**Results:** After data extraction from clinical records, 289 patients had complete data for this assessment, resulting in a 15% of missing values from the initial sample of records reviewed. The baseline prostate volume mean was 47 grams with a standard deviation of 14 grams, and the follow up mean was estimated at 26 grams with a standard deviation of 9 grams. After statistical comparison, this reduction was statistically significant for the observed evidence. A difference between the sample means can be expected to be as extreme as 21 grams with a p-value < 0.0001% of the times under the null hypothesis. Therefore, there is sufficient evidence to reject the null hypothesis. An IPSS of 17,47 score units (6,46 score units of standard deviation) was obtained before treatment for the 289 patient records reviewed, whereas reported IPSS after treatment was estimated with an average of 2score units with a standard deviation of 2 score units. Mean difference contrast for IPSS showed a statistically significant difference with a p-value less than 0,0001, and estimating an average difference of 14,72 score units. A change on the symptom manifestation after treatment is related with a clinical quality of life indicator. Urinary flow rate increased to an average of 19 ml/sec and a standard deviation of 5 ml/sec after procedure was applied. A linear regression model was adjusted using number of applications, age, prostate volume at baseline, IPSS at baseline and urinary flow rate at baseline; obtaining an variability explanation rate of 57% from these variables.

**Conclusion:** There is enough evidence to expect an effective reduction on the prostate volume when applying Rezūm® water vapor therapy in Costa Rican men. IPSS also showed significant evidence to support the efficacy impact on the reduction of symptoms (mean difference was 14,72 average score units). An effect size of 21 grams of difference after applications was obtained with 1.7 standard deviations lower than the baseline measure.

**Interpretation:** A difference in prostate volume of 20.97 average grams was obtained and quality of life was increased significantly, after REZUM treatment was applied.

## 1 Introduction

Benign prostatic hyperplasia is more prevalent in men who have reached their fourth and fifth decades of life [1] and it is also considered to have an impact in patients quality of life [2] reflected in increasing lower urinary tract symptoms(LUTS).

Currently, the standard treatment for treating BPH is performed through surgery that sometimes are invasive and with undesirable and avoidable side effects with the application of minimally invasive therapies such as Rezūm®.

Rezūm® is an ablative minimally invasive treatment for BPH/LUTS that has previously shown beneficial results for patients with BPH diagnosis, thus clinical observational study was conducted with a Costa Rican population, in order to obtain evidence of the potential advantages for a cohort in 2022 [3], [4], [5], [6]. The significance of collecting data in Costa Rican populations was validated with a systematic review that evaluated 11 retrospective studies, 5 prospective observational studies, 2 case series and only one randomized control trial[7]. There are at least 11 sites with different research designs enrolled in the USA, other represented countries are France, United Kingdom, Italy, Ireland and there is one pilot prospective designed cohort enrolled in 3 sites in Dominican Republic, Czech Republic, and Sweden.

In this sense, the evaluation of effectiveness validity of treatments that enable the reversal of benign hyperplasia at the prostate level is very relevant, especially to influence on the quality of life of male population. It is important to note that this impact promises a potential improvement for the population in general, since it has a direct effect on self-esteem and sexual function, upgrading interpersonal relationships and fulfilling functions in different areas of a man’s life.

Therefore the present study undertakes a retrospective observation based on clinical records to obtain a mean difference and a effect size between a basal measure of the prostate volume and a follow up measure. International Prostate Symptom Score (IPSS) was also used to contrast a baseline with a prospective measurement to obtain a contrast of the two observation times [8]. Regression analysis was also estimated in order to explore on the correlations of the studied clinical factors.

## 2 Methods and materials

A retrospective observational study is proposed, based on the review of records and the extraction of variables related to lower urinary tract symptoms. Records reviewed were consulted from the HULI electronic data base of the UNIBE Hospital. Specific data extraction on the measurement of prostate volume before and after the Rezūm® water vapor therapy injection is collected to elaborate the aimed for the estimated effect size.

### 2.1 About Rezūm®

This procedure is carried out under mild sedation in an outpatient setting in the urology consultation office. All patients have been previously screened with negative urine culture and whom prostate cancer has been ruled out as well. After the procedure is completed an 18 French urethral catheter is left in place for 5 days. The time of the entire procedure takes around 15 minutes from the moment they enter the procedure office to the moment they exit the room. Patients are then instructed to return to a follow up visit at 1, 3 and 6 months. In these consultation, IPSS, urinary rate and prostate size are recorded via trans abdominal ultrasound with a 3.5Hz convex probe.

### 2.2 Sources of information

The main parameter of this analysis is based on the measure of the prostate volume. Firstly, the volume is observed when the patient attends to urology clinical consultancy, this observation time is considered the baseline of the study. After application, the patient is followed up for up to three months, when another measurement of the prostate volume is collected. This procedure involves the use of a 3.5 hz convex abdominal ultrasound probe with a full bladder or at least holding around 200cc of urine.

Records are going to be reviewed to extract data from dates of intervention, patient’s age at intervention and the following clinical parameters:

− Baseline prostate volume: Weight in grams of the prostate at the time of diagnosis. In this case, a discrete random variable is composed with a minimum value of 10 grams and a maximum value of 100 at a theoretical level.
− Follow up prostate volume: Weight in grams of the prostate after the application of Rezūm® water vapor therapy. In this case, a discrete random variable is composed with a minimum value of 10 grams and a maximum value of 100 at a theoretical level.
− IPSS before and after the procedure: Evaluation of the frequency of symptoms reported by the person at the time of diagnosis and after the application of therapy. It is made up of a 5-point Likert scale where 0 equals the symptom never occurring, 1 refers to a frequency less than 1 time in 5, 2 equals a frequency less than half of the time, 3 refers to half the time, 4 equals more than half the time, and a value of 5 refers to almost always. This scale includes 7 types of prostate symptoms such as:

**Incomplete emptying** and answers the question: How many times have you had the sensation of not completely emptying your bladder after urinating?,

**Frequency** and means that after urinating, how often did you have to urinate again in less than 2 hours?,

**Intermittency** answering the question How often have you noticed that your urinary flow stops and comes back several times while you urinate?,

**Urgency** and that answers the question: How often have you had difficulty holding back the urge to urinate?,

**Weak urinary flow** answering the question How often have you had a weak urinary flow?,

**Effort** and answer the question: How often have you had to strain or strain to start urinating? and

**Nocturia** answering the question: How many times do you get up to urinate from the time you go to bed at night until you get up in the morning?

− Urinary flow rate before and after the procedure: It refers to the maximum value level of urine reached by the flow during micturitions. It is measure in ml per second.

### 2.3 Statistical analysis

A descriptive model estimation process was performed to contrast a significant difference and effect size of the treatment.

Observations were synthesized as mean values and standard deviations were calculated to elaborate a confidence interval for each parameter estimation of the present study.

Distribution of the variable for the measurement of prostate volume is also considered for this analysis.

#### Model specification

Here we define how the estimation function is addressing the research problem of the present study. Statistical techniques are applied to estimate the mean difference between two measures taken to the same participant.

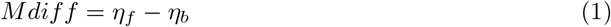

Where *Mdiff* represents the mean difference, *η* is the prostate volume participants distribution, *f* refers to the follow up measurement in the same individual as the *b* distribution. This second distribution *b* represents the baseline prostate volume.

In order to estimate an effect size, the Cohen’s *d* was calculated, then the previous equation can be formulated as follows:

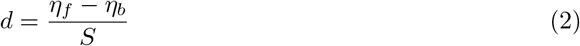

Where *d* is the effect size and *S* represents the standard deviation, assuming both measures have equal variability. This assumption is going to be tested and sensitivity analysis is going to be performed with alternative estimations of the effect size as Hedges’s *g* and Glass’s *δ* to allow selection of the best parameter estimation according to the evaluated data distributions.

A linear regression analysis is also going to be assessed to obtain correlations between the estimated follow up prostate volume and variables like age, baseline prostate volume, IPSS and urinary rate.

Mean difference is estimated with two different time distributions of prostate volume for each of the participants. Baseline prostate volume represents the time before the application of Rezūm® treatment and a follow up measurement of the prostate volume after this applications. Therefore, mean difference is obtained after substraction of this two distributions. A standard deviation will also be calculated to estimate a confidence interval with 95% of probability that the mean value to be considered between the two limits at this level of density. Distribution parameters will also be estimated stratifying the age variable with an interval of 10years for each age category. This mean difference will estimate the analytical efficacy of the treatment. Parameter estimation is undertaken using STATA software. Excel is also used to manage data and typeset tables and specific estimations of the parameter variability.

The obtained model with the previous research design mentioned above, is interpretative as the efficacy of Rezūm® with this sample of participants.

Additionally, the analysis by age is also of relevance in order to identify particular elevated prostate volumes for some or one age group category. This provides information about the state of the volume at the time when participants decide to attend for clinical consultancy.

## 3 Results and discussion

There were three hundred forty three (343) records reviewed of men that had received a Rezūm® intervention to treat their BPH condition. These men were 64 years old in average with a standard deviation of 9 years old. However, there was a case that did not have any information for the variables of interest like baseline prostate volume and it was excluded, providing an cohort of 342 registered men for this analysis. Average baseline prostate volume of these initial cohort was 47.15 grams and had a standard deviation of 14.56 grams.

Average baseline IPSS was estimated at 17 points of the scale with a standard deviation of 6 points and average urinary flow rate was calculated at 12 ml/sec with a standard deviation of 4 ml/sec. These two observations of IPSS and urinary flow rate presented a very low proportion of missing values, where the baseline IPSS had 3% (11) of missing values, the urinary flow rate at baseline had and increased proportion of 7% (23) of cases with no data collected.

Variables collected for the observation of follow up after Rezūm® was applied presented an increased proportion of missing values. For the follow up prostate volume there were 289 cases collected, for a missing value proportion of 15% (53 cases). For the observation of the follow up IPSS, there were 50 cases with missing data representing a proportion of 14%. The urinary flow rate had a missing value proportion of 17% with 58 cases that were not collected for this analysis. Missing data at the moment, refers to cases that was not able to undertake screening after the intervention, because patients did not return for the follow up appointment, as they left Costa Rica and just a few cases of death were presented according to the clinical records included. Therefore, we have an effective cohort of 289 cases that have completed data for the follow up prostate volume and the baseline measures, 53 initial cases were excluded for the analysis. These missing value proportions do not represent a significant reduction of the evidence and estimations were generated with the unbalanced data, specially of the urinary flow rate measures. No data imputation was undertaken.

For a better understanding of the data included, we first are going to characterize these case exclusions. Average age was estimated around 65years old with an standard deviation of 10 years. Average baseline prostate volume was 47 grams with a standard deviation of 17 grams. They had 7 applications of the water vapor injections with a standard deviation of 1application, however applications were registered for only 14 of the 53 patients excluded. The IPSS average at baseline was 17 points with a standard deviation of 7 points and was collected for 42 cases and IPSS values measured at follow up appointments presented an average of 3 points of the scale with a standard deviation of 2 points, however this second value of the IPSS was collected only in 4 cases. In relation to urinary flow rate, 32 patients presented a measure for this variable and average was 14 ml/sec with a standard deviation of 4 ml/sec. We only have one patient record for this group at follow up, showing an urinary flow rate increase to 23 ml/sec from 14 ml/sec.

It is important to annotate, that average prostate volume from the excluded cases at baseline presents a difference of 0.62 grams when it is compared with the 289 cases included. Difference in standard deviation is estimated at 3.17 grams, showing that the included cases have less variability for this observation. In this sense, there is no significant difference between the two groups (p=0.804562).

In Figure 1, the distribution of the baseline prostate volume and the follow up measure of the prostate volume can be compared. These distributions show that after application of Rezūm® water vapor therapies intervention the prostate volume is reduced for the majority of the cases reviewed. The linear graph of this figure is sorted by age, so there is an appreciation of the uniform effect across all ages. There was only one case that presented a reversed volume effect. This patient had a follow up prostate volume of 70 grams after an initial volume of 50 grams. Even though, the case presented an IPSS change of 13 points, showing a considerable difference of the prostate symptoms experience for this patient of 68 years old.

**Fig. 1:**
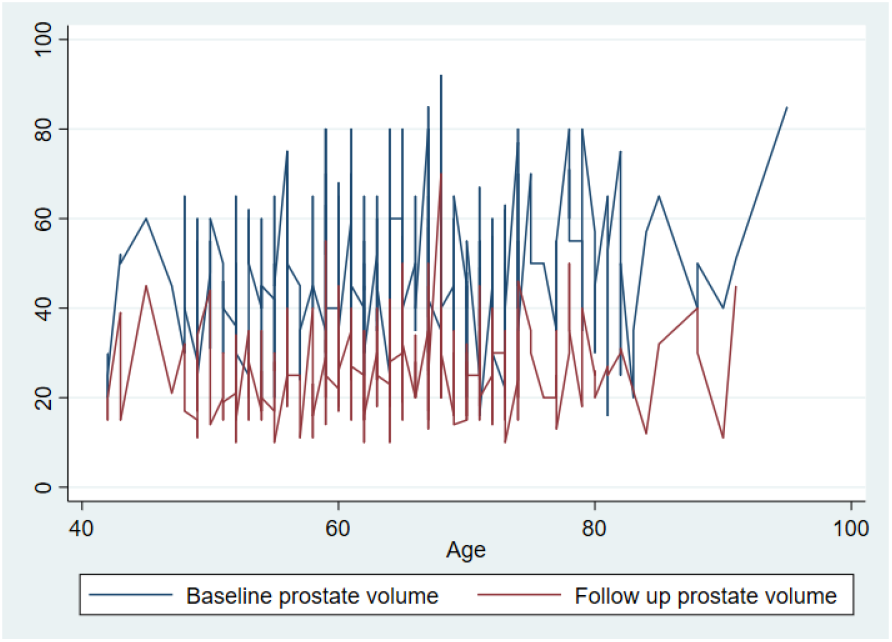
Line distributions of the prostate volume measured in grams, at baseline and three months of follow up after application of Rezūm® water vapor procedures, according to age of the patient.

Patient interventions were practice from December 2020 with 6 records identified. In 2021 there were 136 patients intervened, in 2022 there were 106 patients and in 2023 41 patients were included in this analysis.

For a closer view of the scale, in figure 2 a histogram of prostate volume observed at baseline is drawn and in figure 3 a second histogram is presented for the follow up prostate volume. Firstly, three groups can be identified by a three modal distribution in both histograms. In figure 2 the highest prostate volume density is between 40 grams and 45 grams (frequency of 40 cases with 40 grams and 30 cases with 45 grams), second highest density is located at 50 grams (frequency of 50 cases) and the third mode present 80 grams (frequency of 13 cases). It can be observed that the scale of the prostate volume is centered between 40 grams and 50 grams.

**Fig. 2:**
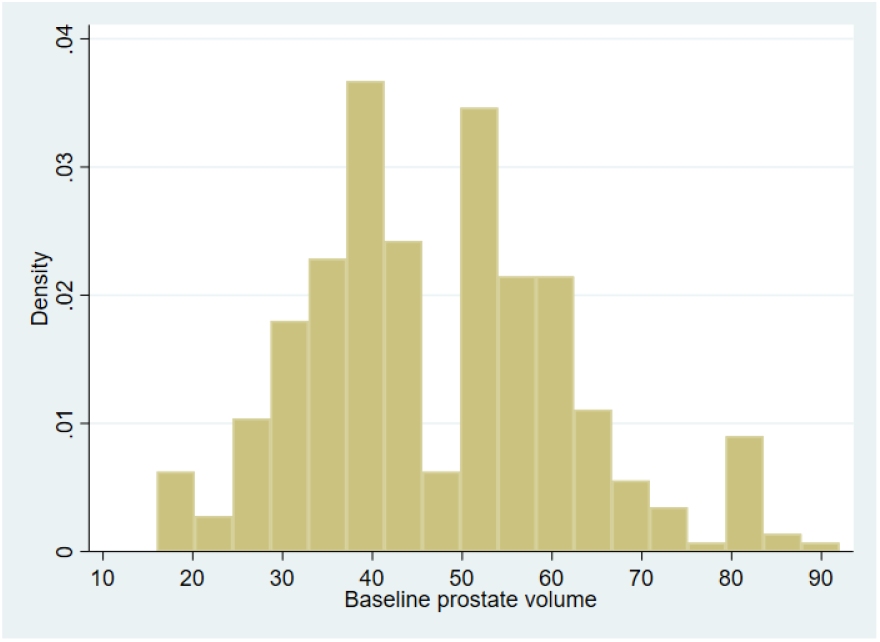
Histogram of prostate volume measured in grams at baseline. Patients with Benign Hyperplasia diagnosis.

**Fig. 3:**
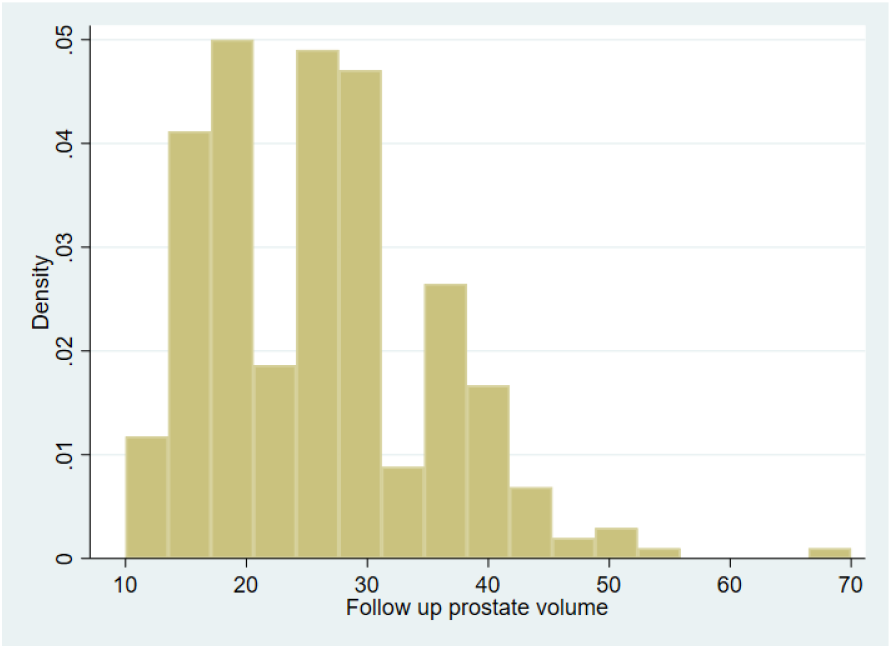
Histogram of prostate volume in grams after application of Rezūm® water vapor therapies. Measure taken after three months of follow up.

Instead in figure 3, the scale is shifted to the left of the axis and the three modes are also reduced. There are 41 cases with a prostate volume of 20 grams, followed by 36 cases with a volume of 25 grams, 34 cases with a volume of 30 grams, 25 cases of 35 grams of volume and 15 cases of 40 grams. After this last group of 40 grams cases decrease considerably, in comparison with the figure 2, this group represented the first highest frequency of the prostate volume. Whereas, after the procedure application relevant modes are between 20 grams and 30 grams.

In figure 4, a comparison of the prostate volume distributions before and after the procedure are visualized. Blue box averages is located at the largest interval limit of the red box, meaning the average of the baseline prostate volume can be reduced at least one standard deviation. Follow up red box present considerable less variability with a standard deviation of 9 grams compared to the baseline standard deviation of 14 grams. The difference between the two variability parameters can offer a precision interpretation of the procedure as the distribution gets more compact and confidence interval limits are closer to the box, even though there are more outliers, these are also under the values of the baseline distribution.

**Fig. 4:**
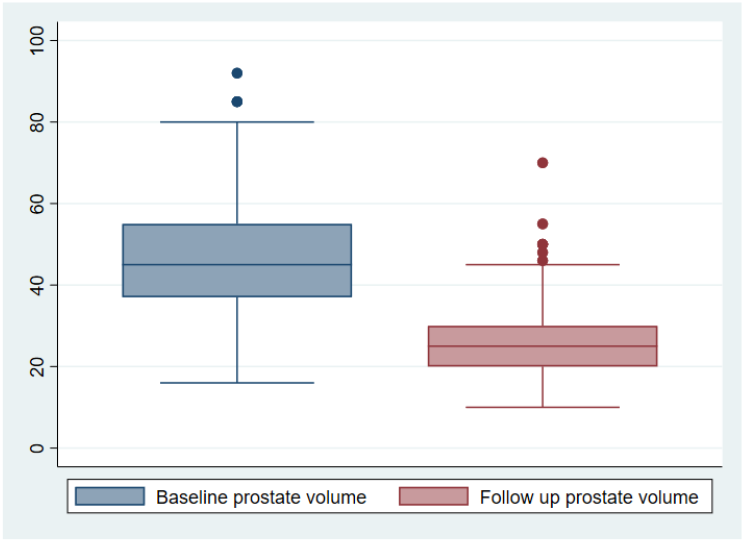
Comparative box plot between prostate volume at baseline of the benign hyperplasia and after three months of follow up of the intervention whith Rezūm® water vapor therapies.

In order to understand differences before and after water vapor injections, age categories were defined by decades to have a better look of the distribution by age. Frequency distributions for each age group were 6% (20 cases) in the 40 to 49 decade, 24% (85 cases) for the 50 to 59 decade, 43% (148 cases) for the 60 to 69 decade, 4% (16 cases) for the 80 to 89 decade and 0.87% (3 cases) for the 90 to 99 decade; where 95 was the maximum age recorded.

In figure 5 six groups of box plots are presented for each of the age groups defined. It can be observed that the group of 60 years to 69 years is the one that display more variability for the prostate volume distribution. However, each of the groups appear to be considerably different.

**Fig. 5:**
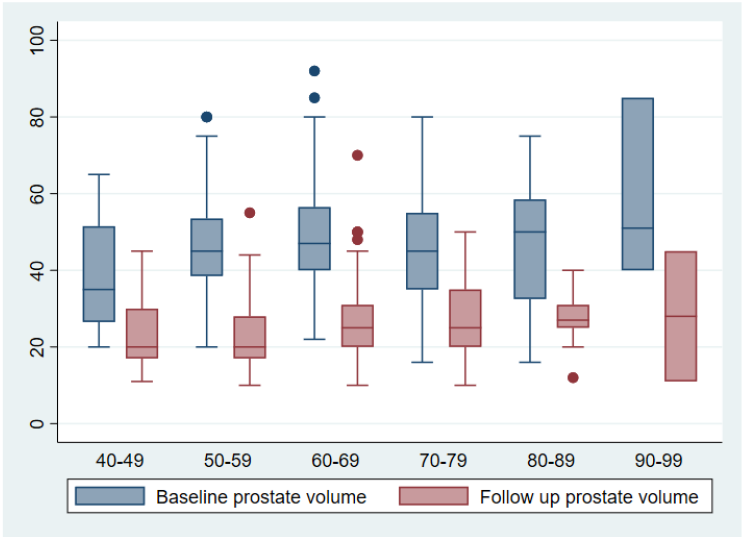
Comparative box plot of prostate volume between baseline benign hyperplasia and after three months of Rezūm® water vapor applications follow up, according to age categories.

Average difference between the two time measures (before and after procedure) 21 grams and had a standard deviation of 10 grams. The two sample z-test applied for the mean difference hypothesis contrast estimated a p-value = 0.0000, showing a significant difference between the baseline parameter of observation and the volume in grams after procedure applications.

The different effect sizes estimated showed a convergence between the Cohen’s *d* and the Hedges’s *g*. First effect size estimated a value of 1.708 grams with a 95% confidence interval of 1.52 grams in the lowest limit and 1.89 grams in the highest limit. For the second effect size parameter estimation the value was 1.706 grams with a 95% confidence interval of 1.52 grams to 1.88 grams. These estimates indicate that follow up prostate volume is around 1.7 standard deviations lower than the baseline prostate volumes. The estimates using Glass’s *δ* 1 reported an effect size of 1.45grams with a 95% confidence interval of 1.25 grams to 1.64 grams and the Glass’s *δ* 2 estimated an effect size of 2.31 with a 95% confidence interval of 2.06 grams for the lowest limit and 2.55 grams for the highest. This parameter shows that the comparison of two times of the prostate volume can reach up to 2 standard deviations of difference after the procedure is applied.

To incorporate the IPSS and the urinary flow rate into the analysis of the effect size before and after the intervention, a two sample *z* test was applied to each of the variables. For the IPSS the parameter contrast was also significant with a differential estimate of 14 points of decrease in the prostate symptom scale used, with a p-value equals to 0.0000. In the same direction of contrast was obtained for the urinary flow rate, estimating a differential rate of -7.49 ml/sec and a p-value equals to 0.0000. In the case of the urinary flow rate the expectation is an increased observation in consequence of the reduction of the prostate there is more space for the bladder to regulate urine.

In figure 6 the differential IPSS distribution by age categories can be detailed. An average of 17 points and a standard deviation of 6points was obtained at baseline. Therefore, let interpret the reduction in the IPSS scale resulting in a uniform distribution with an average of 2 points and a standard deviation of 2 points. It can be observed that the IPSS distribution increases its point count by age at baseline. The highest variability can be identified for the central decades between 50 years old to 79 years old. It is also important that there is a relevant change in the quality of life of the registered men specially at a higher age, nonetheless this is the less represented age group.

**Fig. 6:**
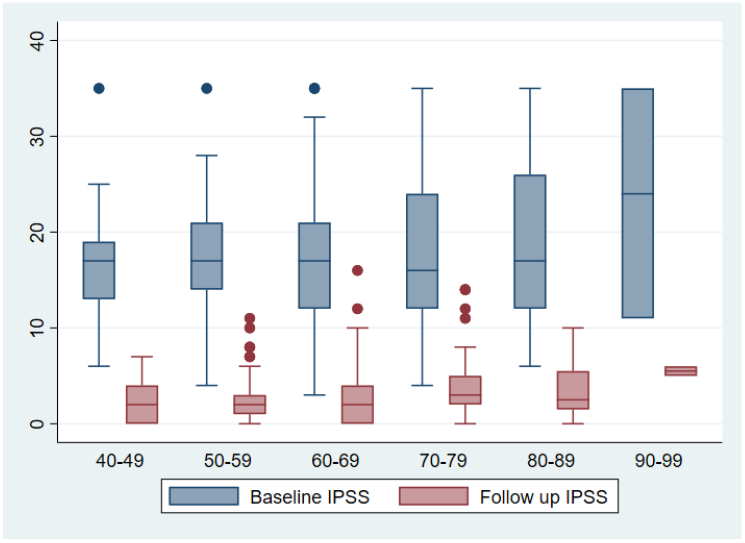
Comparative box plot of International Prostate Symptom Score distribution at baseline when the patient is diagnosed with Benign Hyperplasia and after Rezūm® water vapor intervention with three months of follow up, according to age categories.

When the urinary flow rate is examined, the scales have a change because this parameter tends to increase when the prostate volume is reduced. In figure 7, the parameter distribution can be observed for each box plot estimated by age. At baseline the minimum urinary flow rate was 4 ml/sec and the maximum was 26 ml/sec, whereas after treatment the minimum reached 10 ml/sec and the maximum capacity observed 47 ml/sec. Average urinary flow rate at baseline was 12 ml/sec with a standard deviation of 4 ml/sec and after the procedure the average went up to 19 ml/sec and a standard deviation of 6 ml/sec. This measure presents more variability after the procedure. Age groups with more variability are identified from 50 years old to 69 years old.

**Fig. 7:**
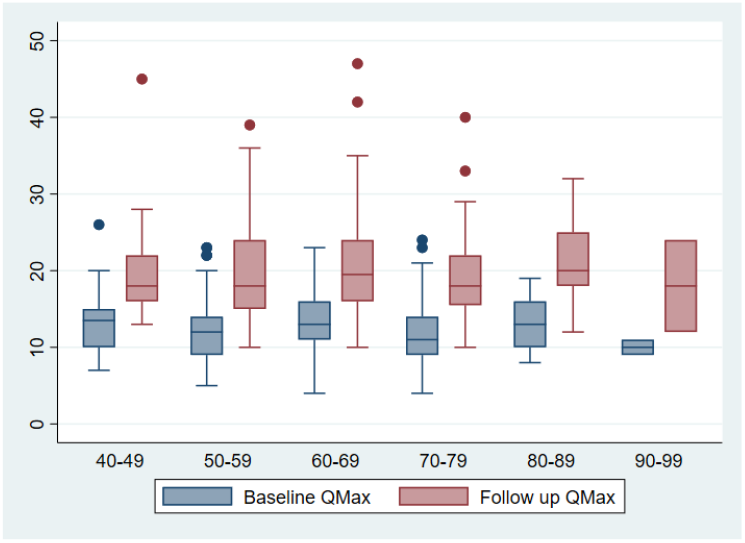
Comparative box plot of Urinary Flow rate (Qmax) distribution at baseline of the Benign Hyperplasia diagnosis and after Rezūm® water vapor therapies with three months of follow up, according to age categories.

**Fig. 8:**
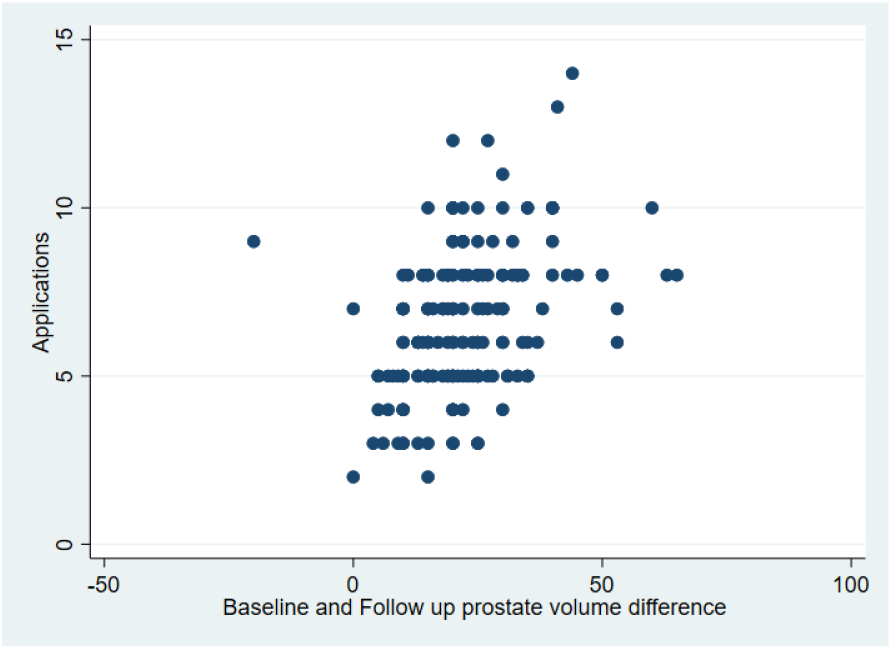
Scatter plot of differential values (horizontal axis) of the substraction between prostate volume when the Benign Hyperplasia is diagnosed and after application of Rezūm® water vapor therapies with three months of follow up. The vertical values are representing the number of applications the patients had received during the intervention.

The number of applications was also included in the analysis to identify the relation with the reduction of the prostate. However, this variable was the one with more missing values as only 167 records were obtained for the included patients.

A Pearson correlation between the difference or value of reduction and the number of applications is 0.42 and is significant at a level of 0.05 with a p-value of *p <* 0.00001. This correlation indicates that at a higher level of applications more reduction is probable to reach. The mean number of applications was 6 times, with an estimated standard deviation of 2 times and a minimum of 2 applications and a maximum of 14 applications.

A mean difference was estimated at 21 grams with a standard deviation of 10 grams after application of Rezūm®.

A linear regression model was adjusted for the estimation of the applications using the follow up prostate volume as a response variable. This model calculated an determination correlation of 8%. Therefore, after different models were estimated the model with previous variables describe in this section were included obtaining the following regression equation:

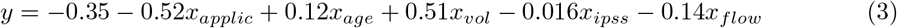

where, *y* **represents the follow up prostate volume**, as we are interested in estimating the prostate volume after procedure is applied; *applic* represents the number of applications when the patient is intervene; *age* represents the age of the patient at the time of the intervention; *vol* represents the volume of the prostate at baseline; *ipss* represents the symptom scale at baseline and flow represents the urinary flow rate at baseline.

This equation means that the number of applications reduces the prostate volume in 0.51 units when this number increases in one application (p=0.036); age increases by 0.12 units of the follow up prostate volume when the age increases in one year (p=0.011); the resulting prostate volume is also affected positively by 0.51 units when the baseline prostate volume is increased by one gram (p=0.000); the IPSS influences the volume after procedure negatively in 0.016 units, if it is increased by one point in the scale (p=0.789); and the urinary flow rate also can influence negatively the prostate volume after by 0.14 units when this measure increases in one ml/sec (p=0.150). This model estimated a determination correlation of 0.59, showing that 59% of the variability of the prostate volume, after Rezūm® applications were conducted, is explained by the included variables in this model. Even when some of the variables are not correlating significantly to the model, we consider that have clinical significance as the results showed above.

Further work of this analysis should include variables related to erectile dysfunction and ejaculation, in order to measure how this factors are influenced after these therapies. Metabolic conditions are also relevant to take into account in the analytical model, to assess how related prostate symptoms can change, for a better understanding on how they are related with BPH.

## 4 Ethical revisions

A protocol was submitted, reviewed and approved by the ethics commitee of CEC UNIBE, and was registered with the code CEC-UNIBE CEC-07-2023. A public registry was made in clinicaltrials.gov, with the code NCT06315062.

## 5 Conclusion

In conclusion, a significant reduction of average 21 grams in prostate volume was observed after a mean of 6 applications in 289 patients. This reduction is reflected in the reduction of prostate symptoms reported by patients. This evidence can be interpreted as an increase in their quality of life after water vapor therapy.

The effect size of Rezūm® to reduce prostate volume is estimated at 1.70 grams (at least one standard deviation of difference) with a 95% confidence interval of 1.52 grams to 1.89 grams. Demonstrating a considerable significant efficacy of this procedure for the treatment of BPH. This means that there is a high probability of BPH reversal after average of 6 applications.

## Data Availability

All relevant data are within the manuscript and its Supporting Information files. If requested data can be available vía email.

## 6 Acknowledgments

This work was made possible thanks to the Nurse Leily Cerdas, as she supported the data extraction from each of the records included. We also want to thank Dr. Dean Elterman for his detailed review and contribution to clear up previous versions of this analysis and scientific report.

## 7 Contributions

Dr. Mario González undertook and directed the design of the study. Dra. Milena Castro performed the statistical analysis for the study. Both authors contributed to the edition and compilation of this manuscript.

## References

1. H. Lepor, “Pathophysiology, epidemiology, and natural history of benign prostatic hyperplasia,” Reviews in urology, vol. 6, no. Suppl 9, p. S3, 2004.

2. E. Fernández-Guzmán, A. A. Matas, V. C. Poves, J. R. Zuazu, P. G. Abad, J. Martínez-Salamanca, L. Q. Franco, J. Justo-Quintas, J. Romero-Otero, and M. Domínguez-Esteban, “Preliminary results of a national multicenter study on the treatment of luts secondary to benign prostatic hyperplasia using the rezūm® steam system,” Actas Urológicas Españolas (English Edition), 2022.

3. C. Alegorides, M. Fourmarier, C. Eghazarian, S. Lebdai, A. Chevrot, and S. Droupy, “Treatment of benign prostate hyperplasia using the rezum® water vapor therapy system: results at one year,” Progrès en Urologie, vol. 30, no. 12, pp. 624–631, 2020.

4. R. F. Tutrone and W. Schiff, “Early patient experience following treatment with the urolift prostatic urethral lift and rezum steam injection,” Can J Urol, vol. 27, no. 3, pp. 10213–10219, 2020.

5. M. J. Johnston, M. Noureldin, Y. Abdelmotagly, L. Paramore, T. Gehring, T. G. Nedas, G. Rajkumar, A. Emara, and R. G. Hindley, “Rezum water vapour therapy: promising early outcomes from the first uk series,” BJU international, vol. 126, no. 5, pp. 557–558, 2020.

6. Z. Green, J. Westwood, and B. K. Somani, “What’s new in rezum: a transurethral water vapour therapy for bph,” Current urology reports, vol. 20, no. 7, pp. 1–7, 2019.

7. M. Babar, J. Loloi, K. Tang, U. Syed, and M. Ciatto, “Emerging outcomes of water vapor thermal therapy (rezum) in a broad range of patients with lower urinary tract symptoms secondary to benign prostatic hyperplasia: A systematic review,” LUTS: Lower Urinary Tract Symptoms, vol. 14, no. 3, pp. 140–154, 2022.

8. J. Bosch, W. Hop, W. Kirkels, and F. Schröder, “The international prostate symptom score in a community-based sample of men between 55 and 74 years of age: prevalence and correlation of symptoms with age, prostate volume, flow rate and residual urine volume,” British journal of urology, vol. 75, no. 5, pp. 622–630, 1995.

